# PREVALENCE AND FACTORS ASSOCIATED WITH FALSE NEGATIVE RAPID DIAGNOSTIC HIV TESTS AMONG PATIENTS ON ANTIRETROVIRAL THERAPY WITH A NON-DETECTABLE VIRAL LOAD IN KAMPALA, UGANDA: A CROSS-SECTIONAL STUDY

**DOI:** 10.64898/2026.02.19.26346688

**Authors:** Nagawa Elizabeth, Lydia Nakiyingi, Joan N. Kalyango, Shivan Nuwasiima, Job Mukwatamundu, Douglas Bulafu, Mikka Brian, Stuart Niwagaba, Ndagga George, Sean Steven Puleh, Patience Muwanguzi, Joaniter I. Nankabirwa

**Affiliations:** Clinical Epidemiology Unit, School of Medicine, Makerere University College of Health Sciences, Kampala, Uganda; Department of Internal Medicine, School of Medicine, Makerere University College of Health Sciences, Kampala, Uganda; Infectious Disease Institute, Makerere University College of Health Sciences, Kampala, Uganda; Department of Pharmacy, School of Health Sciences, Makerere University College of Health Sciences, Kampala, Uganda; Infectious Diseases Research Collaboration (IDRC), Kampala, Uganda; Department of Disease Control and Environmental Health, School of Public Health, Makerere University College of Health Sciences, Kampala, Uganda; MBN Clinical Laboratories; Department of Epidemiology and Biostatistics, Faculty of Public Health, Lira University. Lira, Uganda; Department of Nursing, School of Health Sciences, Makerere University College of Health Sciences, Kampala, Uganda

**Author notes:** **Corresponding author**: Nagawa Elizabeth. P.O BOX 7072, Kampala.

**Keywords:** Prevalence, False-negative HIV result, PLWHIV, Undetectable viral loads

## Abstract

**Background:** Evidence emerging from Sub-Saharan Africa indicates that people living with HIV (PLHIV) on long-term antiretroviral therapy (ART) especially when the viral load is undetectable, may falsely test negative for HIV on rapid diagnostic tests. This study assessed the prevalence and factors associated with false negative rapid diagnostic HIV tests among Patients on antiretroviral therapy, with undetectable viral load levels at Kisenyi Health Center IV, Kampala, Uganda.

**Methods:** Between October 2023 and February 2024, a cross-sectional study was conducted among 1,248 PLHIV on ART with undetectable viral loads at Kisenyi Health Center IV. Participants were recruited consecutively, and HIV re-testing was conducted in accordance with the national serial rapid testing algorithm. The algorithm includes a screening test (Determine™ HIV-1/2), a confirmatory test (Stat-Pak®), and a tie-breaker test (SD Bioline®). Enzyme-linked immunosorbent assay (ELISA) was used as the final confirmatory method. Data on socio-demographics and clinical characteristics was collected using an electronic data abstraction tool. Logistic regression analysis was done to assess for factors associated with false negative results, using STATA version 14.0.

**Results:** The median age of the participants was 34.0 (interquartile range 29.0-42.5 years). The prevalence of false-negative rapid test results was 3.2% (40/1248; CI:2.20-4.2). CD4 (aOR 1.001, CI:1.001-1.003) and duration on ART (aOR 0.884, CI:0.801-0.978) were significantly associated with false-negative HIV results.

**Conclusion:** False-negative results were observed in approximately 3 in every 100 PLHIV on ART with an undetectable viral load. Serial rapid testing alone may be suboptimal for detecting HIV infection in this population. Further confirmatory testing in individuals who test negative on rapid testing is recommended.

## INTRODUCTION

Globally, HIV affected approximately 40.8 million people and accounted for 630 000 deaths in 2024[1]. Sub-Saharan Africa remains the most heavily impacted region, accounting for 26.3 million people living with HIV (PLHIV), representing two-thirds of global burden [1]. In Uganda, the HIV prevalence was 4.9% in 2024, equating to approximately 1.5 million PLHIV [2]. To combat the epidemic, Uganda adopted the World Health Organization (WHO) ’Test and Treat’ strategy in 2016. This approach involves early HIV diagnosis through regular testing of high risk populations and initiating antiretroviral therapy (ART) for all individuals diagnosed with HIV, regardless of CD4 count or clinical stage [3]. With early diagnosis, PLHIV have had prompt ART initiation, leading to reduced viral load, and consequently lower risk of transmission [4].

In most African countries, including Uganda, HIV diagnosis is performed using a serial testing algorithm based on rapid diagnostic tests (RDTs) that detect HIV-1/2 antibodies. The algorithm typically begins with a highly sensitive screening test, followed by a confirmatory antibody-based RDT, and, if necessary, a tie-breaker test, as outlined in the national HIV testing guidelines [5, 6]. However, evidence from several African countries has reported reduced sensitivity of RDTs especially among PLHIV with persistently undetectable viral loads, resulting in false-negative results [7–10]. This is largely attributed to waning HIV antibodies in individuals on long-term ART [9, 11]. These false-negative results are sometimes misinterpreted as evidence of HIV cure by patients, potentially leading to confusion, mismanagement, and even premature discontinuation of ART [12]. Data from the District Health Information System (DHIS), showed that 4,115 PLHIV in Kampala district alone interrupted ART for over three months between October 2020 and September 2021, a trend that may partially reflect this challenge [13].

Although several studies have assessed false-negative HIV results in children and adolescents on ART, adult populations have been relatively underrepresented, in sub-Saharan Africa. In this study, we assessed the prevalence and factors associated with false-negative HIV test results among PLHIV, on ART and with undetectable viral loads. The findings generated aimed to strengthen the evidence base on false HIV negative results, inform policy decisions, and enhance clinical practice among health healthcare workers to prevent unintended treatment interruptions. Addressing this issue is essential to advancing global efforts aimed at ending the HIV epidemic by 2030.

## METHODs

### Study design and setting

A cross-sectional study was conducted at Kisenyi Health Center IV between October 2023 and February 2024. Kisenyi Health Center IV is located in the central division of Kampala district, the capital city of Uganda. Kampala has an HIV prevalence of 6.9%, which is higher than the national average of 4.9% [14]. Kisenyi is one of the ten government-owned health centers under Kampala City Council Authority (KCCA). The health center provides free outpatient and inpatient maternal and outpatient services to predominantly peri-urban and slum populations, and hosts the largest public ART clinic in Kampala. It typically serves between 350-450 PLHIV per week consistent with its classification as a high-volume health facility.

### Study population and sampling

The study consecutively screened PLHIV who were receiving care at Kisenyi ART clinic within the study period. Participants were enrolled if they fulfilled the following criteria: i) aged 18 months and above; ii) on ART; iii) had undetectable viral load and iv) provided informed consent to be part of the study. For children, consent was provided by their caregivers, and assent was obtained from all children aged 8 and above. PLHIV who had interrupted ART in the last one-year, and/or those who were classified as having advanced disease (stage III and IV) were excluded.

### Sample size estimation

The sample size was calculated using the formula for comparison of two proportions [15, 16]. The calculation assumed 5% level of significance, 80% power, and comparison of false negative results among subjects with early and delayed ART initiation. The assumed proportions of subjects with early and delayed ART initiation were 0.87 and 0.13 respectively [11]. In addition, the assumed proportion of false HIV negative results in subjects with an early ART initiation was 0.039 while the proportion of false HIV negative results in subjects with delayed ART initiation was 0.111 [11]. Based on these assumed values, the calculated sample size was 1,016. After adjusting for 10% missing data, the final sample size was 1,117 participants. This sample size was also adequate to determine the prevalence of false HIV negative results based on assumed prevalence of 4.9%, 95% confidence interval, and 5% level of precision using the formula for single proportions [17]. The final sample size was 1248. However, 1,954 patients were assessed for eligibility, anticipating that some would have detectable viral loads and therefore be excluded following the viral load assessment.

### Variables

The outcome variable was a false HIV-negative test result, defined as a sero-negative result obtained through serial rapid HIV testing using Determine, Stat-Pak, and SD Bioline that was subsequently confirmed as HIV-positive by enzyme-linked immunosorbent assay (ELISA).

The independent variables included:

a. Sociodemographic characteristics: age, sex, level of education, marital status, and occupation.
b. Clinical factors: current antiretroviral therapy (ART) regimen, duration on ART, time interval between initial HIV-positive diagnosis and ART initiation, treatment adherence, CD4 cell count, presence of comorbidities, body mass index (BMI), and alcohol and drug use.

### Data collection Laboratory procedures

From each participant, 8 mL of venous blood was collected using EDTA (purple-top) and plain (red-top) vacutainers under standard aseptic procedures. Four milliliters in the EDTA tube were referred to the Central Public Health Laboratory (CPHL) for HIV-1 viral load quantification. The specimen in the plain vacutainer was centrifuged at 3000 rpm for 10 minutes, and approximately 2 mL of serum was separated, aliquoted into sterile cryovials, and stored in labeled cryo-boxes at −80 °C. On return of the viral load results, participant samples with an undetected viral load were subsequently identified and tested using the serial testing algorithm using the Determine®, STAT-PAK®, and SD Bioline® as guided by the National HIV Testing Services algorithm [5]. Negative results were confirmed with a gold standard Enzyme-linked immunosorbent assay (fourth generation ELISA) [18, 19].

### Other data collection procedures and data management

Using a pre-tested electronic data abstraction tool, data on socio-demographic and clinical characteristics including current ART regimen, viral load, CD4 cell count, adherence, comorbidities, and ART duration was extracted from the participants’ files. The data abstraction was implemented on the Open Data Kit (ODK) platform using ODK Collect version 2023.3.0, which had in-built quality control checks. The data was kept on a separate network and only authorized study staff had access to the data during the collection and processing phase.

### Data analysis

Data analysis was done using STATA version 14.0 (Stata Corporation, College Station, TX). Descriptive statistics were used to summarize data for all variables, means with their standard deviations or medians and their inter-quartile ranges (IQR) for numeric data and proportions for categorical data. The primary outcome was a false HIV negative test result which was defined as having negative HIV test results from the serial testing using Determine, Statpak and SD bio line that was confirmed as positive by the gold standard Architect HIV Ag/Ab Combo Assay. It was computed as a proportion with its 95% confidence interval. Logistic regression was used to determine the factors associated with false HIV negative results in both unadjusted and adjusted models. Variables with p-values less than 0.2 in unadjusted models were included in the adjusted models. Odds ratios with their 95% confidence intervals were computed. Statistical interaction in adjusted models was assessed using the likelihood ratio tests, and confounding was assessed by comparing odds ratios from unadjusted and adjusted models. A p-value less than 0.05 was considered statistically significant in the final model.

### Ethics approval and consent to participate

Ethical approval was obtained from the Makerere University School of Medicine Research and Ethics Committee (protocol number; Mak-SOMREC-2023-603). The Permission to conduct the study at Kisenyi Health Centre IV was obtained from KCCA Directorate of public health and the in-charge of the health facility. All study procedures adhered to national ethical guidelines and the principles of the Declaration of Helsinki.

A written informed consent and assent was obtained from research participants before enrolling for the study. For children, the informed consent was given by caregivers and children aged 8 years and above gave assent. All data were de-identified prior to analysis to ensure participant confidentiality.

## RESULTS

### Characteristics of study participants

Between October 2023 and February 2024, 1954 PLHIV were consecutively screened for eligibility to join the study, and 1248(63.9%) with undetectable viral load were enrolled. The reasons for exclusion included no viral load results (n=33, 1.7%), viral load detected (n=649, 33.2%) and hemolyzed samples (n=24, 1.2%) **(Figure 1).** The median age of the participants was 34.0 (IQR 29.0-42.5 years). Majority were females (76.8%), married/living with a partner (58.0%), and employed (77.0%). Most (96.9%) were on TDF/3TC/DTG regimen of ART and median duration on ART was 6 (IQR 4.0-9.0) years **(Table 1).**

**Figure 1:**
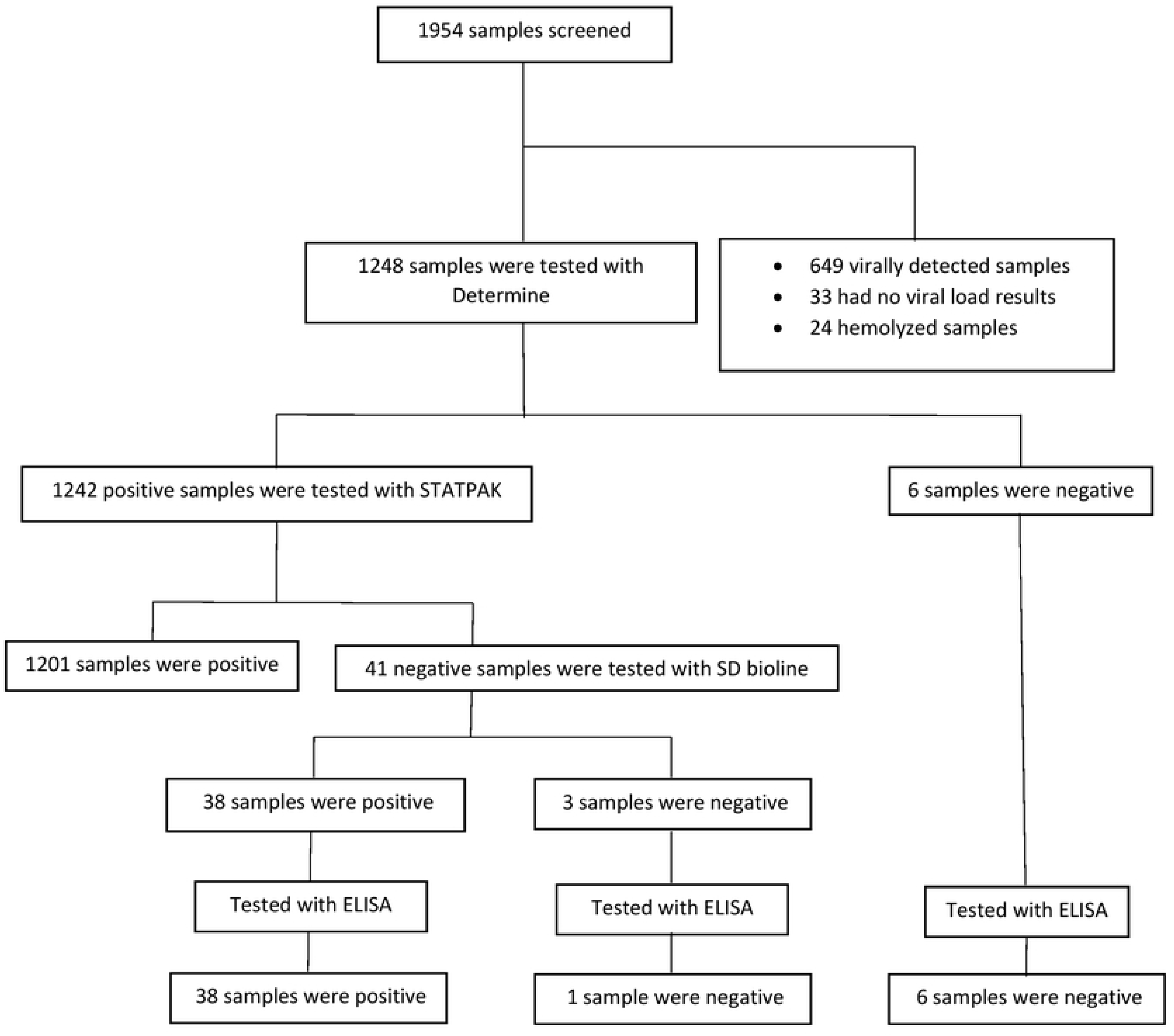
Study Profile of blood samples collected at Kisenyi Health Centre IV, October 2023.

**Table 1:**
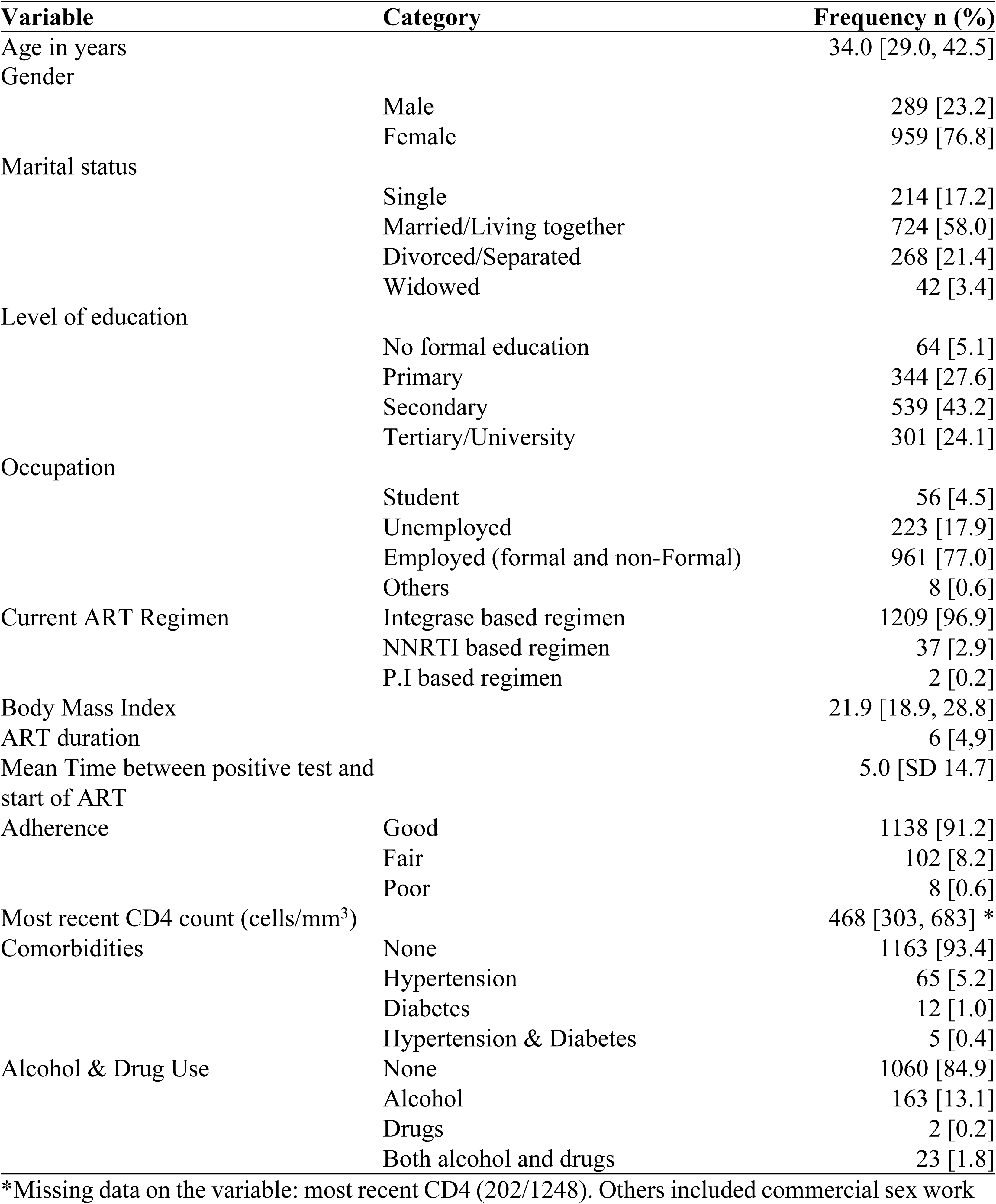
Socio-demographic and clinical characteristics of 1248 study participants at Kisenyi Health Centre IV, October 2023 – February 2024.

### Prevalence of false negative results among 1248 study participants

Among the 1,248 participants, 40 had false-negative rapid HIV results, corresponding to a prevalence of 3.2% (95% CI: 2.18–4.24).

### Unadjusted and adjusted analysis of factors associated with false negative results among 1248 study participants

At unadjusted analysis, age (cOR 0.96 CI: 0.93-0.98, P=0.01), ART duration (cOR 0.92, CI: 0.84-0.99, P=0.052), time between a positive test and the start of treatment (cOR O.89, CI:0.78-0.96, P=0.13), most recent CD4 count (cOR= 1.01, CI:1.00-1.02, P=0.01) had a p-value less than 0.2 and were included in the adjusted analysis. Duration on ART (aOR 0.88, CI:0.80-0.98, P=0.02) and most recent CD4 count (aOR= 1.001, CI:1.001-1.003, P=0.01) were found significant at multivariable analysis. There was no interaction and confounding **(Table 2).**

**Table 2:**
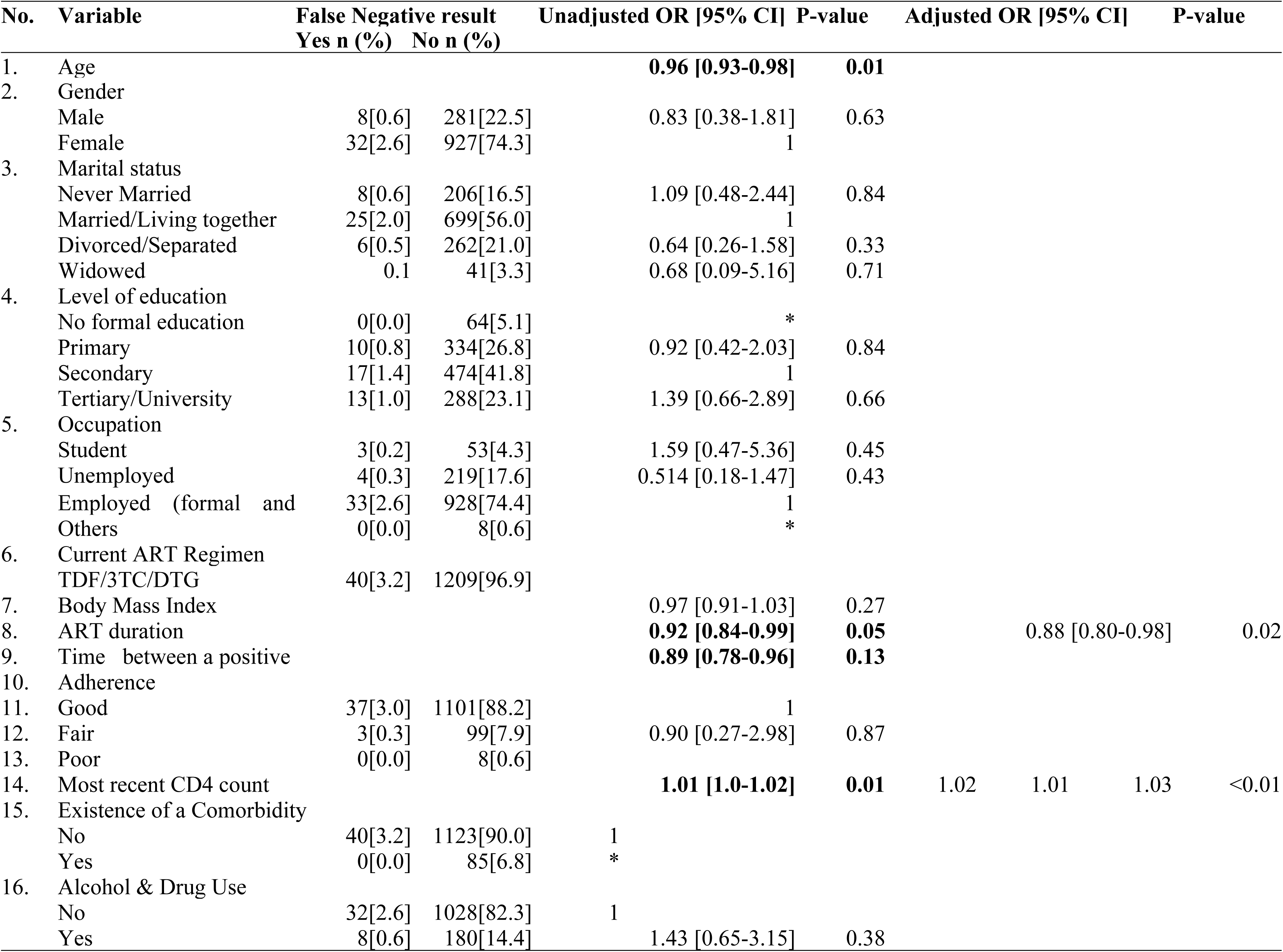
Unadjusted and adjusted analysis of factors associated with false negative results among study participants at Kisenyi Health Centre IV, October 2023 to February 2024.

| No. | Variable | False Negative result |  | Unadjusted OR [95% CI] | P-value | Adjusted OR [95% CI] |  |  | P-value |
| --- | --- | --- | --- | --- | --- | --- | --- | --- | --- |
|  |  | Yes n (%) | No n (%) |  |  |  |  |  |  |
| 1. | Age |  |  | **0.96 [0.93-0.98]** | **0.01** |  |  |  |  |
| 2. | Gender |  |  |  |  |  |  |  |  |
|  | Male | 8[0.6] | 281[22.5] | 0.83 [0.38-1.81] | 0.63 |  |  |  |  |
|  | Female | 32[2.6] | 927[74.3] | 1 |  |  |  |  |  |
| 3. | Marital status |  |  |  |  |  |  |  |  |
|  | Never Married | 8[0.6] | 206[16.5] | 1.09 [0.48-2.44] | 0.84 |  |  |  |  |
|  | Married/Living together | 25[2.0] | 699[56.0] | 1 |  |  |  |  |  |
|  | Divorced/Separated | 6[0.5] | 262[21.0] | 0.64 [0.26-1.58] | 0.33 |  |  |  |  |
|  | Widowed | 0.1 | 41[3.3] | 0.68 [0.09-5.16] | 0.71 |  |  |  |  |
| 4. | Level of education |  |  |  |  |  |  |  |  |
|  | No formal education | 0[0.0] | 64[5.1] | * |  |  |  |  |  |
|  | Primary | 10[0.8] | 334[26.8] | 0.92 [0.42-2.03] | 0.84 |  |  |  |  |
|  | Secondary | 17[1.4] | 474[41.8] | 1 |  |  |  |  |  |
|  | Tertiary/University | 13[1.0] | 288[23.1] | 1.39 [0.66-2.89] | 0.66 |  |  |  |  |
| 5. | Occupation |  |  |  |  |  |  |  |  |
|  | Student | 3[0.2] | 53[4.3] | 1.59 [0.47-5.36] | 0.45 |  |  |  |  |
|  | Unemployed | 4[0.3] | 219[17.6] | 0.514 [0.18-1.47] | 0.43 |  |  |  |  |
|  | Employed (formal and | 33[2.6] | 928[74.4] | 1 |  |  |  |  |  |
|  | Others | 0[0.0] | 8[0.6] | * |  |  |  |  |  |
| 6. | Current ART Regimen |  |  |  |  |  |  |  |  |
|  | TDF/3TC/DTG | 40[3.2] | 1209[96.9] |  |  |  |  |  |  |
| 7. | Body Mass Index |  |  | 0.97 [0.91-1.03] | 0.27 |  |  |  |  |
| 8. | ART duration |  |  | **0.92 [0.84-0.99]** | **0.05** | 0.88 [0.80-0.98] |  |  | 0.02 |
| 9. | Time between a positive |  |  | **0.89 [0.78-0.96]** | **0.13** |  |  |  |  |
| 10. | Adherence |  |  |  |  |  |  |  |  |
| 11. | Good | 37[3.0] | 1101[88.2] | 1 |  |  |  |  |  |
| 12. | Fair | 3[0.3] | 99[7.9] | 0.90 [0.27-2.98] | 0.87 |  |  |  |  |
| 13. | Poor | 0[0.0] | 8[0.6] |  |  |  |  |  |  |
| 14. | Most recent CD4 count |  |  | **1.01 [1.0-1.02]** | **0.01** | 1.02 | 1.01 | 1.03 | <0.01 |
| 15. | Existence of a Comorbidity |  |  |  |  |  |  |  |  |
|  | No | 40[3.2] | 1123[90.0] | 1 |  |  |  |  |  |
|  | Yes | 0[0.0] | 85[6.8] | * |  |  |  |  |  |
| 16. | Alcohol & Drug Use |  |  |  |  |  |  |  |  |
|  | No | 32[2.6] | 1028[82.3] | 1 |  |  |  |  |  |
|  | Yes | 8[0.6] | 180[14.4] | 1.43 [0.65-3.15] | 0.38 |  |  |  |  |

## DISCUSSION

This study evaluated false-negative HIV rapid diagnostic test results among a total of 1248 participants with an undetectable viral load enrolled at Kisenyi Health Centre IV in Kampala, Uganda. All participants were tested for HIV infection using a serial rapid testing algorithm, with ELISA as the gold standard. These findings indicated that 3.2% (95% CI: 2.18–4.24) of negative results from serial rapid testing in people living with HIV (PLHIV) on ART with an undetectable viral load were false negatives, which were confirmed positive by the gold standard ELISA test.

The prevalence of false–negative test results of 3.2% observed in this study aligns closely with results reported in Zimbabwe (3.5%) among 1921 PLHIV, and in Thailand among 234 participants tested using similar protocols [19, 20]. However, slightly higher rates were observed in Belgium (4.9%) among 390 participants [20, 21]. This difference could be explained by variations in populations and HIV testing algorithms, where assays [and/or DNA-PCR] were used as confirmatory tests in the Belgium study [21]. In contrast, a lower prevalence of 2% was found in South Africa [22]. This difference can be explained by the fact that the South African study did not screen participants for viral load status, meaning that individuals with detectable viral loads may have been included in the study. In addition, the variability across settings underscores the importance of local context, particularly viral suppression levels and test kit generations.

False-negative rapid test results among people living with HIV on ART with an undetectable viral load may lead to serious consequences. A negative test may prompt patients to discontinue ART, mistakenly believing they are cured. This can result in viral rebound, increased transmission risk, and progression to advanced disease or death [9, 21]. This finding emphasizes the need for cautious interpretation of negative rapid test results among virally suppressed individuals and reinforces the importance of confirmatory testing, especially when used to verify diagnosis in those previously known to be HIV-positive.

Most recent CD4 cell count was significantly associated with false negative results. A unit increase in CD4 count was associated with a 0.001 increase in the odds of a false HIV negative result. This finding mirrors similar significant results from a longitudinal study conducted in Thailand among 234 participants initiated on ART. It found that a CD4 cell count ≥350 cells/µL increased the likelihood of a false-negative result [20]. Conversely, a cohort study conducted among 207 adults in Malawi and a retrospective study among 268 children in London did not observe a statistically significant association [9, 11]. In the present study, most recent CD4 corresponded to the baseline CD4 count obtained at ART initiation for 59.8% of participants, rather than a follow-up CD4 measured during ART. This may partly explain the observed differences in findings from the studies in Malawi and London that assessed CD4 counts longitudinally during ART follow-up.

ART duration was significantly associated with false negative results where the shorter the duration on ART, the more likely the false negative result could occur. This finding may be attributed to the introduction of DTG based regimens as the preferred first-line regimens in 2018. Notably, all false-negative results in this study were observed among participants who had been initiated on and were currently receiving the DTG based regimens [3]. This may also explain why few children in the study presented with false negative HIV results, as pediatric dolutegravir-based regimens were only introduced in the 2020 Uganda guidelines, with Dolutegravir 10mg becoming available in health facilities in 2022 [23].

In contrast, a study conducted in Belgium among 83 participants found that the length of time on treatment did not have a statistical influence on the probability to obtain a false-negative test result. However, it indicated that the fastest reversion (false HIV negative result) was obtained after 4 months in an early treatment cohort [21]. Among the long-term treated cohort (390 HIV-1 patients with ≥ 9 years of undetectable viral load), longer treatment increased nonreactivity of the HIV rapid tests [21]. These differences could be attributed to variations in study design, treatment regimens, sample size and study populations.

Though age, tertiary education and current ART regimen were significant at bivariate analysis, they became insignificant at multivariate analysis. This could have been due to fact that some variable categories had no/ few numbers of outcome - false negative results.

This study found variables sex, marital status, BMI, viral load, adherence, and time between positive test and ART initiation insignificant. However, studies conducted in Malawi and London found viral load and time between positive test and ART initiation significant [11, 24]. It is worthwhile to note that these studies considered the initial viral load at ART initiation while the current study used the last viral load result on ART follow-up.

### Limitations of the study

The study was conducted at one public health facility located in an urban setting with rigorous readily available government supported HIV care and follow-up; therefore, these findings are not generalizable to all PLHIV, particularly those in private health facilities and rural settings.

Consecutive recruitment of participants could have introduced selection bias. However, this was minimized by enrolling participants on all clinic days, including those attending community Differentiated Service Delivery models (DSDM) and special clinics. This was done at the laboratory, a converging point for all patients due for viral load testing.

Information bias may have been introduced due to adherence assessments based on patient self-reports, while selection bias could have arisen from missing CD4 data (17%, n=212). However, participants with missing CD4 and those with CD4 were not significantly different for variables false negative results, sex, age and current regimen.

## Conclusion

False-negative results on serial testing algorithm occur in 3 persons for every 100 PLHIV on ART with an undetectable viral load. In this study, ART duration and CD4 count were significant predicators. Decreasing ART duration and increasing CD4 count increased the likelihood of false negative results.

Serial rapid testing may not be sufficient in detecting HIV infection among ART patients with an undetectable viral load given the high frequency of false positive results. Therefore, further confirmatory testing is recommended for samples that remained negative. We also recommend longitudinal studies to assess other implicating factors for false negative results such as pre-exposure prophylaxis (PrEP), ART adherence biomarkers, acute/chronic HIV infection, and antibody characteristics among PLHIV.

## Data Availability

STROBE checklist, data collection tools and dataset are available from the Figshare repository https://doi.org/10.6084/m9.figshare.31305526

## Glossary

Sex – defined as binary sex categorization (male/female) that was designated at birth.

## Abbreviations

ART: Antiretroviral Therapy
CD4: Cluster of Differentiation 4
CPHL: Central Public Health Laboratories
DHIS: District Health Information System
DNA-PCR: Deoxyribonucleic acid-Polymerase Chain Reaction
ELISA: Enzyme-Linked Immunosorbent Assay
EMR: Electronic medical records
HIV: Human Immunodeficiency Virus
HTS: HIV testing services
IDI: Infectious Disease Institute
KCCA: Kampala Capital City Authority
MOH: Ministry of Health
ODK: Open data kit
PLHIV: People living with HIV
PrEP: Pre-exposure Prophylaxis
RDT: Rapid diagnostic test
SOMREC: School of Medicine Research and Ethics Committee
TDF/3TC/DTG: Tenofovir/Lamivudine/Dolutegravir
UPHIA: Uganda Population-based HIV Impact Assessment
VL: Viral load

## Acknowledgements

I acknowledge the tireless efforts and good will of CEU lecturers, CEU colleagues, KCCA and staff of the ART clinic, Kisenyi Health Center IV, laboratory staff of AIDS Information Centre, research assistants and family.

## Authors’ Information

Elizabeth Nagawa

Clinical Epidemiology Unit, School of Medicine, Makerere University Uganda.

Lydia Nakiyingi

Department of Medicine, School of Medicine, Makerere University, Kampala, Uganda

Dr. Kalyango Joan

Clinical Epidemiology Unit, School of Medicine, Makerere University, Kampala, Uganda

Shivan Nuwasiima

Clinical Epidemiology Unit, School of Medicine, Makerere University, Kampala, Uganda.

Douglas Bulafu

Department of Disease Control and Environmental Health, School of Public Health, College of Health Sciences, Makerere University. Kampala, Uganda

Job Mukwatamundu

Infectious Diseases Research Collaboration (IDRC), Kampala, Uganda.

Mikka Brian

Infectious Disease Institute, Kampala, Uganda.

Stuart Niwagaba

Infectious Diseases Research Collaboration (IDRC), Kampala, Uganda.

Ndagga George

MBN Clinical Laboratories

Sean Steven Puleh

Department of Epidemiology and Biostatistics, Faculty of Public Health, Lira University. Lira, Uganda.

Patience Muwanguzi

School of Health Sciences, College of Health Sciences, Makerere University, Kampala, Uganda

Joaniter Nankabirwa

Clinical Epidemiology Unit, School of Medicine, Makerere University Uganda.

## Author’s contributions

NE, LN, JN, KJ: Conceptualization, Methodology. NE, JM, NG: Investigation. NE: resources. NE, LN, JN, KJ: Writing - Original Draft. NE, LN, JN, KJ, SN, DB, NG, SSP, PM: Writing - Review & Editing. NE, SN: Data Curation and Visualization. NE, LN, JN, KJ: Supervision and Project administration. All authors read and approved the final manuscript.

## Supporting Information

Dataset https://doi.org/10.6084/m9.figshare.31305526

Laboratory result sheet https://doi.org/10.6084/m9.figshare.31305526

Data abstraction tool https://doi.org/10.6084/m9.figshare.31305526

STROBE checklist https://doi.org/10.6084/m9.figshare.31305526

